# The association between academic pressure and adolescent mental health problems: A systematic review

**DOI:** 10.1101/2023.01.24.23284938

**Authors:** Thomas Steare, Carolina Gutiérrez Muñoz, Alice Sullivan, Gemma Lewis

## Abstract

**Background:** Academic pressure is a potential contributor to adolescent mental health problems, but international evidence on this association has never been synthesised.

**Methods:** We conducted the first systematic review on the association between academic pressure and adolescent depression, anxiety, self-harm, suicidality, suicide attempts and suicide. We searched MEDLINE, PsycINFO, ERIC and Web of Science (core collection) up to November 24, 2022, for studies of school-going children or adolescents, which measured academic pressure or timing within the school year as the exposure and depression, anxiety, self-harm, or suicidal ideation, attempts or suicide as outcomes. Risk of bias was assessed using the Mixed Methods Appraisal Tool. We used narrative synthesis to summarise the evidence. The review was prospectively registered with PROSPERO (CRD42021232702).

**Results:** We included 52 studies primarily from Asia (n=26) and Europe (n=20). Most studies assessed mixed anxiety and depressive symptoms (n=20) or depressive symptoms (n=19). Forty-eight studies found evidence of a positive association between academic pressure or timing within the school year and at least one mental health outcome.

**Limitations:** Most studies were cross-sectional (n=39), adjusted for a narrow range of confounders or had other limitations which limited the strength of causal inferences.

**Conclusions:** We found evidence that academic pressure is a potential candidate for public health interventions which could prevent adolescent mental health problems. Large population-based cohort studies are needed to investigate whether academic pressure is a causal risk factor that should be targeted in school- and policy-based interventions.

**Funding:** UCL Health of the Public; Wellcome Institutional Strategic Support Fund.

## Introduction

Depression and anxiety are the two most common mental health problems, and they often begin during adolescence (Solmi et al., 2021). Non-suicidal self-harm (NSSH) is also common among adolescents and often occurs alongside depression and anxiety (Lundh et al., 2011). Together, these mental health problems are leading risk factors for suicidal ideation, suicide attempts, and suicide (Castellví et al., 2017; Gili et al., 2019).

In many countries, including the UK, there is evidence that rates of depression, anxiety, self-harm and suicide are rising among adolescents(Kõlves and de Leo, 2016; Rodway et al., 2020; Sadler et al., 2018). This rise was apparent before the COVID-19 pandemic which has led to a further increase in depressive symptoms among adolescents (Mansfield et al., 2022). Identifying modifiable risk factors for these mental health problems could inform public health interventions to reduce their rising incidence. Schools are a potential setting for public health interventions that would reach most adolescents. There is some evidence that school-based interventions improve mental health, but many are unsuccessful,(Bonell et al., 2018; Shinde et al., 2018; Šouláková et al., 2019; Weare and Nind, 2011) and we need a better understanding of which risk factors to target.

One common source of stress for adolescents is academic pressure. There is evidence that levels of academic pressure have risen among adolescents in recent decades (Löfstedt et al., 2020). In two large surveys (Fildes et al., 2014; YoungMinds, 2019) adolescents cited academic pressure as one of the top influences on their mental health. In a large study of adolescent suicides in the UK, academic pressure was one of the most reported antecedents in coroner investigations (Rodway et al., 2020).

There is also evidence that teachers and parents are concerned about rising levels of academic pressure and the potential association with adolescent mental health problems (National Education Union, 2019, 2018). Academic pressure is likely to be a broad and multifaceted construct, and there is no generally used definition. Consistent with existing studies, our Patient and Public Involvement work with adolescents and teachers has suggested that academic pressure can be defined as fear of failure, concerns about the future, chronic stress about workload and exams, worries about parental expectations, and competition with peers for grades (Shahmohammadi, 2011; Sun et al., 2011). Academic pressure can therefore be distinguished from transient experiences of test/exam anxiety, which resolve during the short-term. Our PPI work also suggested that academic pressure is influenced by individuals, schools, families, policy, and society. If academic pressure is a potential causal risk factor for adolescent mental health problems, it could be modified through interventions aimed at each of these levels, including universal whole-school approaches.

One meta-analysis of psychosocial risk factors for mental health problems among secondary school students in China found that higher levels of academic pressure were associated with increased depressive symptoms (Tang et al., 2020). However, this study only investigated depressive symptoms and the findings are unlikely to generalise to other countries due to vast differences in educational systems.

To our knowledge, no study has synthesised evidence on the association between academic pressure and a broad range of adolescent mental health outcomes across countries. We conducted the first systematic review investigating the association between academic pressure and adolescent depression, anxiety, self-harm and suicide.

## Methods

This systematic literature review was carried out in accordance with the PRISMA guidelines (Page et al., 2021) The protocol was prospectively registered on PROSPERO, number CRD42021232702.

### Eligibility criteria

We included studies published in English that met the following criteria: Participants:

Children and adolescents attending primary or secondary schools, academies, and sixth-form colleges or their international equivalents. We included state, independent or special educational needs schools. Studies were included if the mean age of the sample was younger than 18 years old (the typical end of school age across many countries).

#### Exposures

We included studies reporting any measure of academic pressure or academic stress. We included studies that measured single or multiple components of academic pressure. Studies were included if they used timing within the year as exposures (considered to be a proxy measure of academic pressure). We excluded studies that only considered test anxiety, as we assumed that this referred to transient anxiety experienced during evaluative testing only, rather than prolonged concerns and stress regarding academic performance or demands linked to the academic pressures of school (Zeidner, 2020).

#### Outcomes

We included studies that reported at least one of the following as outcomes: depression, anxiety, mixed anxiety and depression, NSSH, suicide attempts, suicidal ideation (thoughts or plans), and suicide. Studies were included if these outcomes were ascertained from validated scales, clinical diagnoses, mental health service use or official records, and were self-reported by the adolescent or reported by teachers, parents or clinicians.

### Search strategy

We searched four electronic databases: Medline (OVID) (between 1946 and Feb 19, 2021), PsycINFO (OVID) (between 1806 and Feb 19, 2021), ERIC (ProQuest) (between 1966 and Feb 19, 2021) and Web of Science Core Collection (between 1900 and Feb 19, 2021). We used a combination of keyword and subject heading searches. Full search terms are available in the appendix. There were no limits on publication date.

We supplemented the search strategy with a backwards reference search of included studies, and a forward citation search using Web of Science Core Collection (carried out by TS on 10^th^ June 2021). We ran an updated search of the four electronic databases (on 24^th^ November 2022) to identify eligible studies published since the original search.

### Study selection

Titles and abstracts were screened against eligibility criteria using the Rayyan systematic review web application (Ouzzani et al., 2016). TS screened all titles and abstracts, and CGM independently screened 10%. A third senior author (GL) independently resolved disagreements.

TS screened all papers at full text and CGM double-screened 50. GL resolved disagreements at this stage.

During full-text screening, published measures of academic pressure were inspected, when available, to ascertain whether scales and items were consistent with our definition and therefore met our inclusion criteria. We contacted the corresponding authors of seven studies via email for additional information about their measure of academic pressure. None responded, and no follow-up emails were sent. Four academic pressure scales were sent to translators so the items could be assessed in English.

### Data extraction

Data were extracted by TS using an Excel-based form. We extracted citation details, country, sample size, population type, study design, study setting, and the academic pressure measure used. We extracted all reported effect sizes, effect estimates and confidence intervals from analyses aiming to identify the association between academic pressure and each mental health outcome. To check for accuracy CGM extracted data from 10 included papers. GL resolved 6 disagreements. We contacted the corresponding authors of two studies via email for greater clarity on the outcome data, but they did not respond and no follow-up emails were sent.

### Quality assessment

Study quality was assessed by TS using the Mixed Methods Appraisal Tool (MMAT) (Hong et al., 2018). The MMAT allows quality assessment of quantitative non-randomised study designs including cohort, case-control, and cross-sectional studies, and of quantitative descriptive studies. Quality was assessed according to sample representativeness, the appropriateness of the exposure and outcome measurements, the completeness of outcome data (we considered outcome data to be sufficiently complete if available for 80% of the sample or more), whether confounders were appropriately accounted for, and whether the exposure occurred as intended.

### Data analysis

We conducted a narrative synthesis to describe and summarise the characteristics and results of the included studies. Studies were grouped according to mental health outcomes, categorised as: depression, anxiety, mixed anxiety and depression (including psychosomatic symptoms), mental health service use, NSSH, suicide attempt, suicidal ideation and suicide. Extracted study data from each outcome group was tabulated to aid the synthesis. We did not group by school type as initially planned because we identified no studies with samples solely from primary schools. We reported unadjusted and adjusted effect estimates from each study. We synthesised measures of academic pressure according to the type of scale (specific academic pressure measures, subscales from broader scales, and single items).

Narrative synthesis was led by TS with regular discussion with the research team. We did not conduct a meta-analysis due to the diversity of study designs and measures of academic pressure and mental health outcomes used across studies, limiting the ability to produce meaningful summary estimates of effect (Campbell et al., 2020).

## Results

The PRISMA flow diagram is displayed in Figure 1. We identified 13,020 records from our database searches. Following removal of duplicates, we screened 9,898 papers at title and abstract level, and 106 papers at full text. GL resolved 11 disagreements at title and abstract screening, and 5 at full-text screening. After full-text screening of 89 papers, we included 25 articles reporting on 24 original studies. Seventeen full-text articles could not be retrieved or were published in a language other than English. Backwards and forwards citation searching of included studies yielded 19 extra studies. Two studies that did not appear in the database or citation searches were also included by the review team. The update search identified 1,609 studies. Twenty-seven full texts were screened, and eight articles reporting on seven original studies met eligibility criteria. Backwards and forwards citation searching of the included studies from the update search did not yield any further studies.

**Figure 1.**
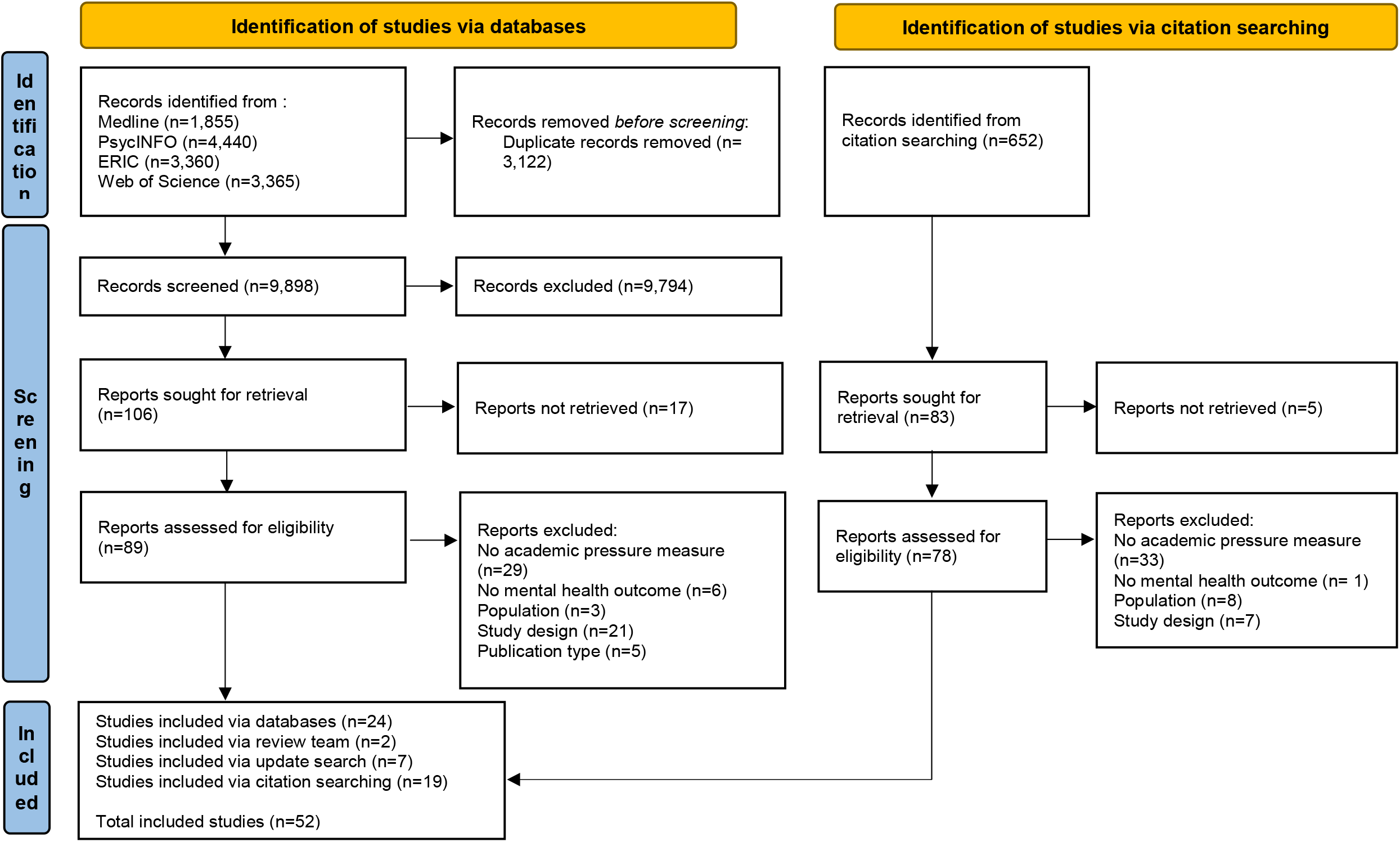
PRISMA flowchart of included studies.

Characteristics of the 52 included studies are displayed in Table 1. Twenty-six were conducted in Asia, 18 in Europe, five in North America, one in Australia, one across Europe and North America, and one across Europe and Australia. Studies were published between 1991 and 2022; 40 from 2010 onwards, suggesting an increased interest in this area.

**Table 1.** Characteristics of included studies.

| Reference | Country | Study design | Participants at follow-up (%) | Sample size | Sample Age (Mean) | Gender (% female) | Academic pressure measure | Outcome category |
| --- | --- | --- | --- | --- | --- | --- | --- | --- |
| (Ang and Huan, 2006a) | Singapore | Cross-sectional | NA | 1,108 | 14.33 | 45.8 | AESI (Ang and Huan, 2006b) | Depression; Suicide thoughts and behaviours |
| (Bersia et al., 2022) | Italy | Repeated cross-sectional | NA | 165,000 |  |  | HBSC single schoolwork pressure item | Mixed depression and anxiety |
| (Blackburn et al., 2021) | UK | Time series | NA | 571,388 |  |  | Timing within year | Service use |
| (Carbone et al., 2019) | USA | Time series | NA | 874,872 |  | 64 | Timing within year | Suicide thoughts and behaviours |
| (Chyu and Chen, 2022) | Hong Kong | Cross-sectional | NA | 1,084 |  |  | 17 item unnamed scale | Mixed depression and anxiety |
| (Cosma et al., 2020) | European countries & Canada | Repeated cross-sectional | NA | 915,054 | 13.54 | 50.8 | HBSC single schoolwork pressure item | Mixed depression and anxiety |
| (Deb et al., 2015) | India | Cross-sectional | NA | 190 | 16.72 | 74.2 | Perception about Stress of Adolescents scale | Mixed depression and anxiety |
| (Eriksson and Sellström, 2010) | Sweden | Repeated cross-sectional | NA | 8,341 |  | 50 | 3 item HBSC scale | Mixed depression and anxiety |
| (Fu et al., 2022) | China | Prospective cohort |  | 6,566 |  | 49.7 | Single parental expectations item | Depression |
| (Guo et al., 2014) | China | Cross-sectional | NA | 1,774 | 16 | 44 | ERI-S (Li et al., 2010) | Depression |
| (Hansen and Lang, 2011) | USA | Time series | NA |  |  |  | Timing within year | Suicide thoughts and behaviours |
| (Hanspal et al., 2019) | India | Cross-sectional | NA | 223 | 12.84 | 42.6 | Single parental pressure item | Depression |
| (Haugan et al., 2021) | Norway | Cross-sectional | NA | 1,077 |  | 56.2 | 3 item HBSC scale | Mixed depression and anxiety |
| (Hawton et al., 2003) | UK | Time series | NA | 1,583 |  | 77.6 | Timing within year | Non-suicidal self-harm |
| (Ho et al., 2022) | Vietnam | Cross-sectional | NA | 1,336 | 15.43 | 53.7 | ESSA (Sun et al., 2011) | Depression |
| (Hodge et al., 1997) | Australia | Cross-sectional | NA | 445 |  | 50.3 | 4 items on examination expectations | Mixed depression and anxiety |
| (Högberg et al., 2020) | Sweden | Repeated cross-sectional | NA | 29,199 | 13.02 | 50.2 | HBSC single schoolwork pressure item | Mixed depression and anxiety |
| (Högberg, 2021) | European countries | Repeated cross-sectional | NA | ≈160,000 | 15.53 | 51 | HBSC single schoolwork pressure item | Mixed depression and anxiety |
| (Hosseinkhani et al., 2020a) | Iran | Cross-sectional | NA | 1724 | 15 | 50.1 | IAASQ (future concerns, academic competition, and stress of parent involvement subscales) (Hosseinkhani et al., | Mixed depression and anxiety |
|  |  |  |  |  |  |  | 2020b) |  |
| (Huang and Chen, 2015) | Taiwan | Cross-sectional | NA | 1,196 | 16.5 | 53.2 | AESI (Chinese translation) (Ang and Huan, 2006b) | Depression |
| (Zahir Izuan et al., 2018) | Malaysia | Cross-sectional | NA | 567 |  | 54.5 | ESSA (Sun et al., 2011) | Mixed depression and anxiety |
| (Jayanthi et al., 2015) | India | Case-control | NA | 1,120 |  |  | ESSA (modified – 20 items) (Sun et al., 2011) | Depression |
| (Jiang et al., 2021)/(Li et al., 2022) | China | Cross-sectional | NA | 552 | 14.48 | 52.5 | Unnamed 4-item scale | Depression; Anxiety |
| (Kaman et al., 2021) | Germany | Prospective cohort |  | 632 | 14.34 | 56 | HBSC single schoolwork pressure item | Depression |
| (Lahti et al., 2007) | Finland | Time series | NA | 25 |  |  | Timing within year | Suicide thoughts and behaviours |
| iu and Lu, 2012) | China | Cross-sectional | NA | 368 | 16.75 | 60 | Academic stress questionnaire (homework subscale) | Depression |
| (Locker and Cropley, 2004) | UK | Prospective cohort | 79.5 | 654 | 14.7 | 40.4 | Proximity to exams | Depression; Anxiety |
| (Long et al., 2020) | UK | Cross-sectional | NA | 2,571 | 15.47 | 50.8 | Single perceived exam pressure item | Mixed depression and anxiety |
| (Lönnfjörð and Hagquist, 2021) | Sweden | Cross-sectional | NA | 2,004 |  | 52 | Adapted schoolwork pressure scale | Mixed depression and anxiety |
| (Ma et al., 2018) | Hong Kong | Cross-sectional | NA | 872 | 15.65 |  | HPE (Fuligni, 1997) | Depression |
| (Masood et al., 2018) | Pakistan | Case-control | NA | 200 | 15.04 | 50 | ESSA (Sun et al., 2011) | Suicide thoughts and behaviours |
| (Matsubayashi et al., 2016) | Japan | Time series | NA | 14,071 |  |  | Timing within year | Suicide thoughts and behaviours |
| (McCleary et al., 1991) | USA | Time series | NA |  |  |  | Timing within year | Suicide thoughts and behaviours |
| (Moksnes et al., 2016a, 2016b) | Norway | Cross-sectional | NA | 1,289 | 15 | 51.2 | ASQ-N (school performance subscale) (Moksnes et al., 2010) | Depression |
| (Nguyen et al., 2013) | Vietnam | Cross-sectional | NA | 1,161 | 16.1 | 63.5 | ESSA (Sun et al., 2011) | Depression; Anxiety |
| (Nygren and Hagquist, 2019) | Sweden | Repeated cross-sectional | NA | 20,152 |  |  | HBSC single school demands item | Mixed depression and anxiety |
| (Quach et al., 2015) | China | Cross-sectional | NA | 997 |  | 52 | IPI (academic pressure subscale) (Campbell, 1994) | Depression; Anxiety |
| (Redmond et al., 2022) | Australia, England & Spain | Cross-sectional | NA | 11,200 |  |  | HBSC single schoolwork pressure item | Mixed depression and anxiety |
| (Ringdal et al., 2020) | Norway | Cross-sectional | NA | 1,814 |  | 51.7 | ASQ-N (school performance subscale) (Moksnes et al., 2010) | Mixed depression and anxiety |
| (Shang et al., 2014) | China | Cross-sectional | NA | 1,004 | 15.8 | 53.4 | ERI-S (Li et al., 2010) | Suicide thoughts and behaviours |
| (Slaunwhite et al., 2019) | Canada | Time series | NA | 4,410 | 15.83 | 57.3 | Timing within year | Service use |
| (Song et al., 2019) | China | Cross-sectional | NA | 411 | 13.4 | 48.7 | Effort-Reward Imbalance for Learning Scale (Fukuda et al., 2010) | Depression |
| (Song et al., 2020) | China | Cross-sectional | NA | 5,959 | 11.56 | 47.7 | ASLEC (Liu et al., 1997) | Mixed depression and anxiety |
| (Sonmark et al., 2016) | France & Sweden | Repeated cross-sectional | NA | 21,467 |  |  | HBSC single schoolwork pressure item | Mixed depression and anxiety |
| (Spiller et al., 2020) | USA | Time series | NA | 922,930 |  | 76.8 | Timing within year | Suicide thoughts and behaviours |
| (Sun and Hui, 2007) | Hong Kong | Cross-sectional | NA | 1,358 | 13.8 | 49.9 | Chinese Academic Pressure Scale | Depression; Suicide thoughts and behaviours |
| (Torsheim and Wold, 2001) | Norway | Cross-sectional | NA | 1,585 | 13.5 |  | 3 item HBSC scale | Mixed depression and anxiety |
| (Torsheim et al., 2003) | Norway | Prospective cohort | 77.8 | 767 | 13.9 |  | HBSC Academic Stress subscale + High Academic Expectations subscale | Mixed depression and anxiety |
| (Wahab et al., 2013) | Malaysia | Cross-sectional | NA | 350 | 16 | 39.4 | 3SQ (Academic related stressor subscale) (Yusoff, 2011) | Depression; Anxiety |
| (Wen et al., 2020) | China | Cross-sectional | NA | 900 | 14.14 |  | Single academic pressure item | Anxiety |
| (Zhang et al., 2013) | China | Cross-sectional | NA | 1,297 |  | 47.3 | ASLEC (Liu et al., 1997) | Depression |
| (Zhang et al., 2019) | China | Cross-sectional | NA | 33,635 |  |  | Single academic pressure item | Suicide thoughts and behaviours |

AESI = Academic Expectations Stress Inventory; ASLEC = Adolescent Self-Rating Life Events Checklist; ASQ-N = Norwegian version of the Adolescent Stress Questionnaire; ESSA = Educational Stress Scale for Adolescents; ERI-S = Effort-Reward Imbalance at School; HBSC = Health Behaviour in School-aged Children; HPE = High Parental Expectations; IAASQ = Iranian Adolescents Academic Stress Questionnaire; IPI = Inventory of Parental Influence; 3SQ = Secondary School Stressor Questionnaire. NA = not applicable.

Most studies were cross-sectional (n=39), with 33 using data from one time-point, and six using repeated cross-sectional surveys at multiple time-points. Two cross-sectional studies used a case-control design (Jayanthi et al., 2015; Masood et al., 2018). We found three prospective cohort studies, which measured academic pressure at baseline and mental health at follow-up (follow-up ranged from one to five years) (Fu et al., 2022; Kaman et al., 2021; Torsheim et al., 2003). A further prospective cohort study investigated how mental health outcomes differed according to proximity to examinations (Locker and Cropley, 2004) We found nine longitudinal studies that used a time series design, where data were collected from mental health services or administrative records, to assess associations between mental health service use or outcomes and timing within the year (Blackburn et al., 2021; Carbone et al., 2019; Hansen and Lang, 2011; Hawton et al., 2003; Lahti et al., 2007; Matsubayashi et al., 2016; McCleary et al., 1991; Slaunwhite et al., 2019; Spiller et al., 2020). Sample sizes were generally large, with 34 studies containing over 1,000 participants. The sample sizes for (a) cross-sectional and repeated cross-sectional, (b) case-control (c) prospective cohort, and (d) time-series designs ranged from 190 to 915,054; 200 to 1,120; 632 to 6,566; and 25 to 922,930 respectively.

Thirty-three studies sampled from secondary schools, three from both primary and secondary schools, and 16 studies did not state the school type. Across studies, the mean age of participants ranged from 11.5 to 16.8. Academic pressure was measured using a scale or sub-scale in 29 studies, and with a single item in 13 studies. Ten studies used timing within the year or proximity to examinations as the exposure.

### Risk of bias

The risk of bias varied across different domains of the MMAT (Table S1; appendix pp 14-15). Thirty-two studies were identified as having a sample representative of the target population. Nineteen studies provided insufficient detail on the target population, inclusion criteria and reasons for non-participation to be rated. Thirty-one studies were rated as using appropriate measures. Twenty-one studies were rated low in quality as we were unable to find evidence of the development or validation of the academic pressure measure. Twenty-two studies reported complete outcome data, with 28 providing no details on missing data or attrition. Only two studies reported that outcome data were obtained from fewer than 80% of participants. Twenty-eight studies were judged to sufficiently account for confounders. All studies were rated as academic pressure occurring as expected. Overall, nine included studies were not rated as at risk of bias across all the MMAT domains.

### Academic pressure measures

Twenty-seven different single-items or scales were used to measure academic pressure, all self-reported by students (Table S2; appendix pp 16-20). Measures varied in the aspects of academic pressure assessed, but most used items on stress or pressure from school, schoolwork, exams, or parents. Only a few measures assessed other aspects of academic pressure, such as concerns about future prospects following school, school failure, and competitiveness with peers or siblings.

We found 16 scales specifically developed to assess academic pressure, used in 22 studies. We found evidence of sufficient item development, scale development, and validation in independent samples for four scales (Boateng et al., 2018). The Academic Expectations Stress Inventory (AESI) is a 9-item scale that measures academic stress owing to expectations from parents, teachers, and the self (Ang and Huan, 2006b). The scale has demonstrated acceptable reliability and validity in samples of adolescents attending secondary schools in Singapore, and was used in two studies (Ang and Huan, 2006a; Huang and Chen, 2015). The most common measure was the Educational Stress Scale for Adolescents (ESSA) (n=5) (Sun et al., 2011). The ESSA uses adapted items from the AESI to assess academic stress across five domains: pressure from study, workload, worry about grades, self-expectation, and despondency. The ESSA was originally tested and developed in a large sample of Chinese adolescents and has demonstrated acceptable reliability and internal consistency. We found two Chinese scales that assessed academic stress through the imbalance of effort and reward in school or learning (Effort-Reward Imbalance for Learning Scale; Effort–Reward Imbalance at School), both showing acceptable validity and reliability (Fukuda et al., 2010; Li et al., 2010).

The High Parental Expectations (HPE) four-item scale, developed in a study of American students at high and middle schools, was used in one study (Ma et al., 2018). The HPE scale assesses perceived parental pressure towards academic achievement and attainment and displayed good internal consistency in the original study (Fuligni, 1997). We found no evidence of scale development or validation.

Other measures of academic pressure included scales developed specifically for the study they were used in (Chyu and Chen, 2022; Deb et al., 2015b; Eriksson and Sellström, 2010; Fu et al., 2022; Haugan et al., 2021; Hodge et al., 1997; Jiang et al., 2021; Li et al., 2022; Lönnfjord and Hagquist, 2021; Sun and Hui, 2007; Torsheim et al., 2003; Torsheim and Wold, 2001). We were unable to identify evidence of development of these measures. In comparison to the validated scales these typically featured fewer items and a narrow focus on single aspects of academic pressure such as schoolwork pressure or examination expectations.

We found five academic pressure subscales within broader measures of adolescent stress, which were used in seven studies. These subscales included “Study Pressure” from the Adolescent Self-Rating Life Events Checklist (n=2) (Liu et al., 1997), “School Performance” from the Norwegian version of the Adolescent Stress Questionnaire (n=2) (Moksnes et al., 2010), “Academic-related stressors” from the Secondary School Stressor Questionnaire (n=1) (Yusoff, 2011) “Academic Pressure stressors” from the Inventory of Parental Influence (n=1)(Campbell, 1994), and “Future Concerns”, “Academic Competition”, and “Stress of Parent Involvement” from the Iranian Adolescents Academic Stress Questionnaire (n=1) (Hosseinkhani et al., 2020b).

We identified seven single item measures used in 13 studies, which assessed level of pressure or stress from exams, parents, schoolwork, teachers, or school.

### Narrative synthesis

#### Depression

Depression was the second-most common outcome, with 19 studies included (Table 2). Depression outcome measures included depressive symptoms (n=15), approximations of clinical diagnoses (n=3), and a latent construct of psychological adjustment comprising depressive symptoms, loneliness, and self-esteem (n=1) (Song et al., 2019) Sixteen cross-sectional studies reported positive associations between academic pressure and depression, with one finding no evidence of an association. In one case-control study, adolescents that met the criteria for depression reported greater levels of academic stress than controls (Jayanthi et al., 2015).

**Table 2.**
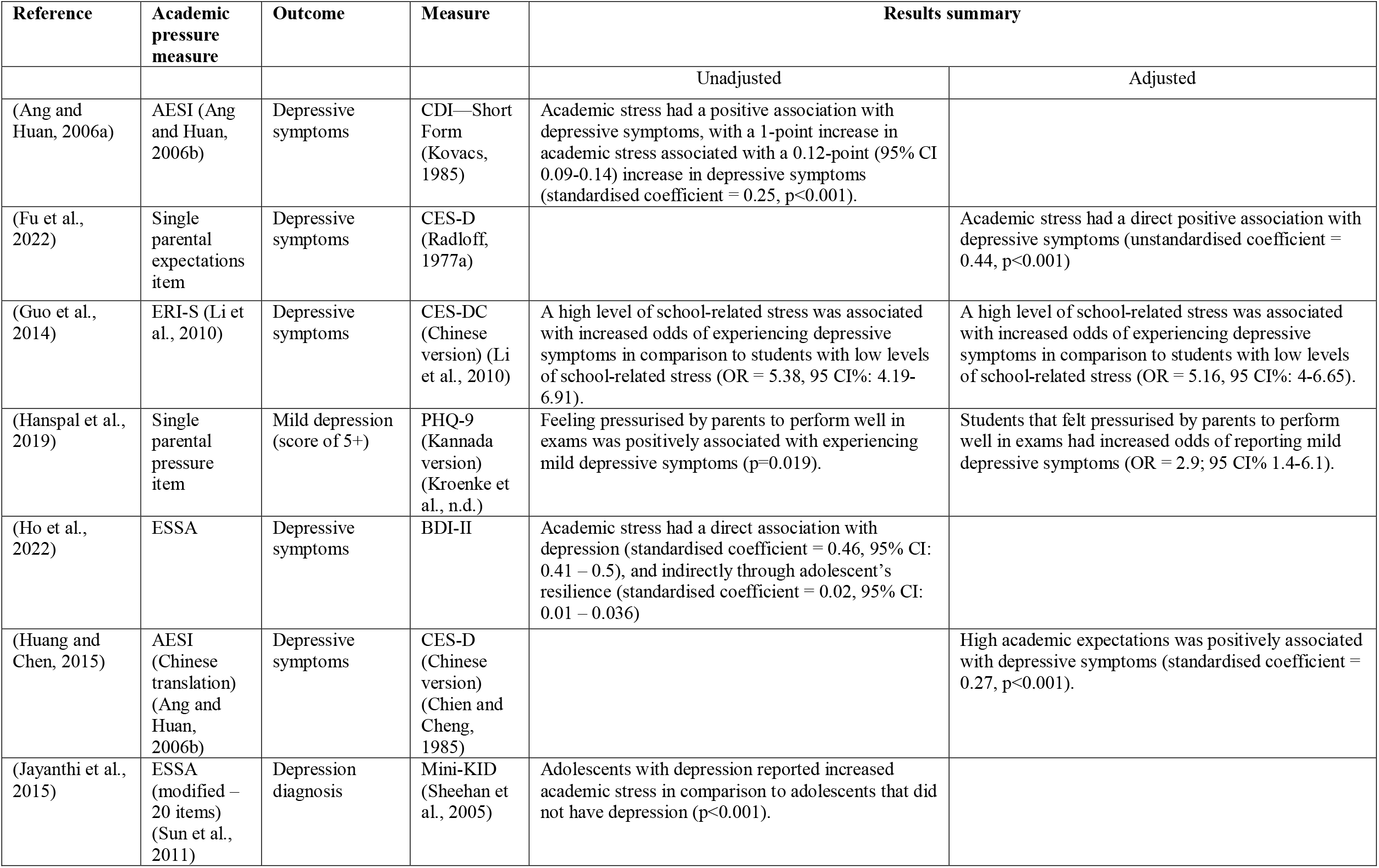
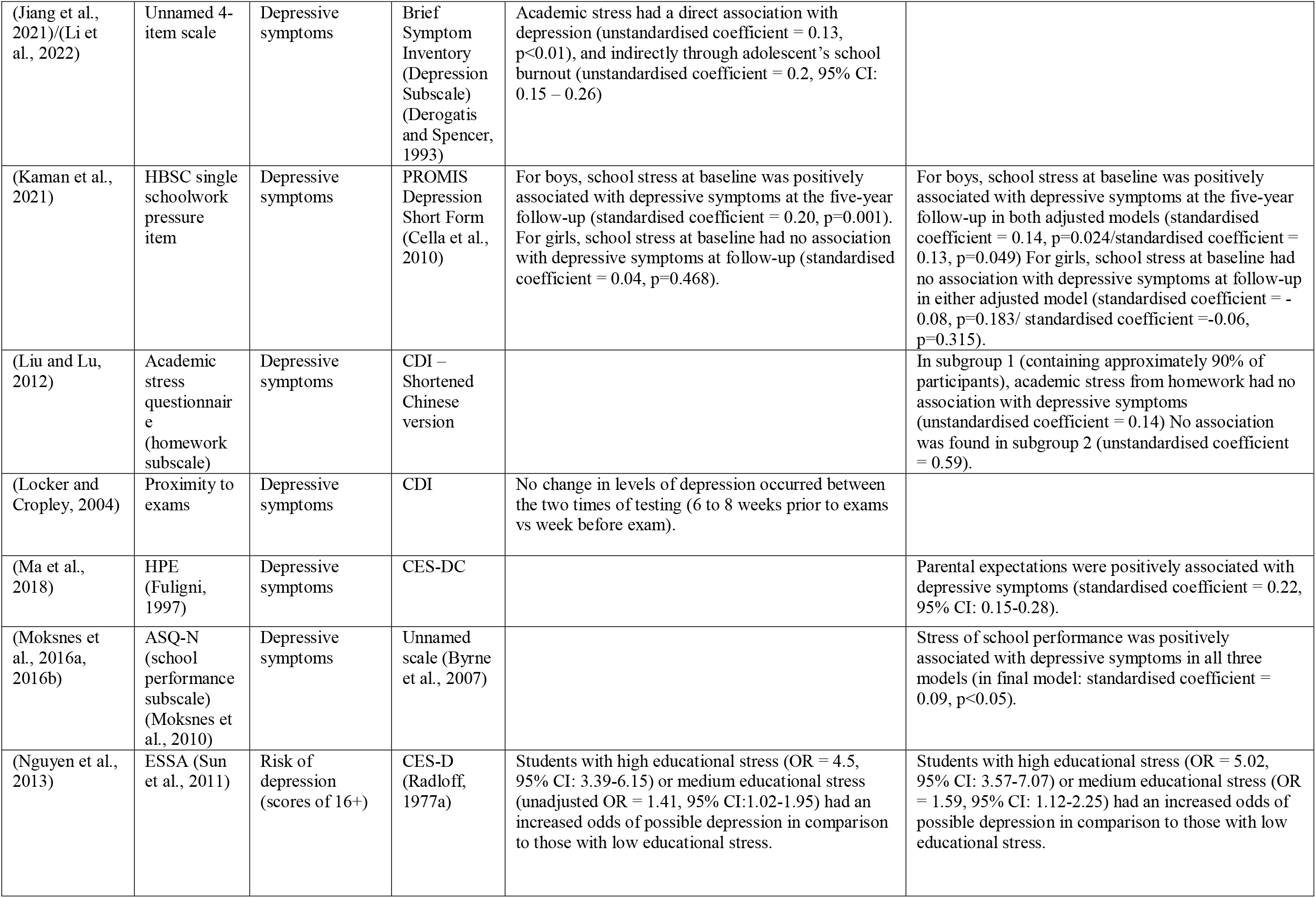
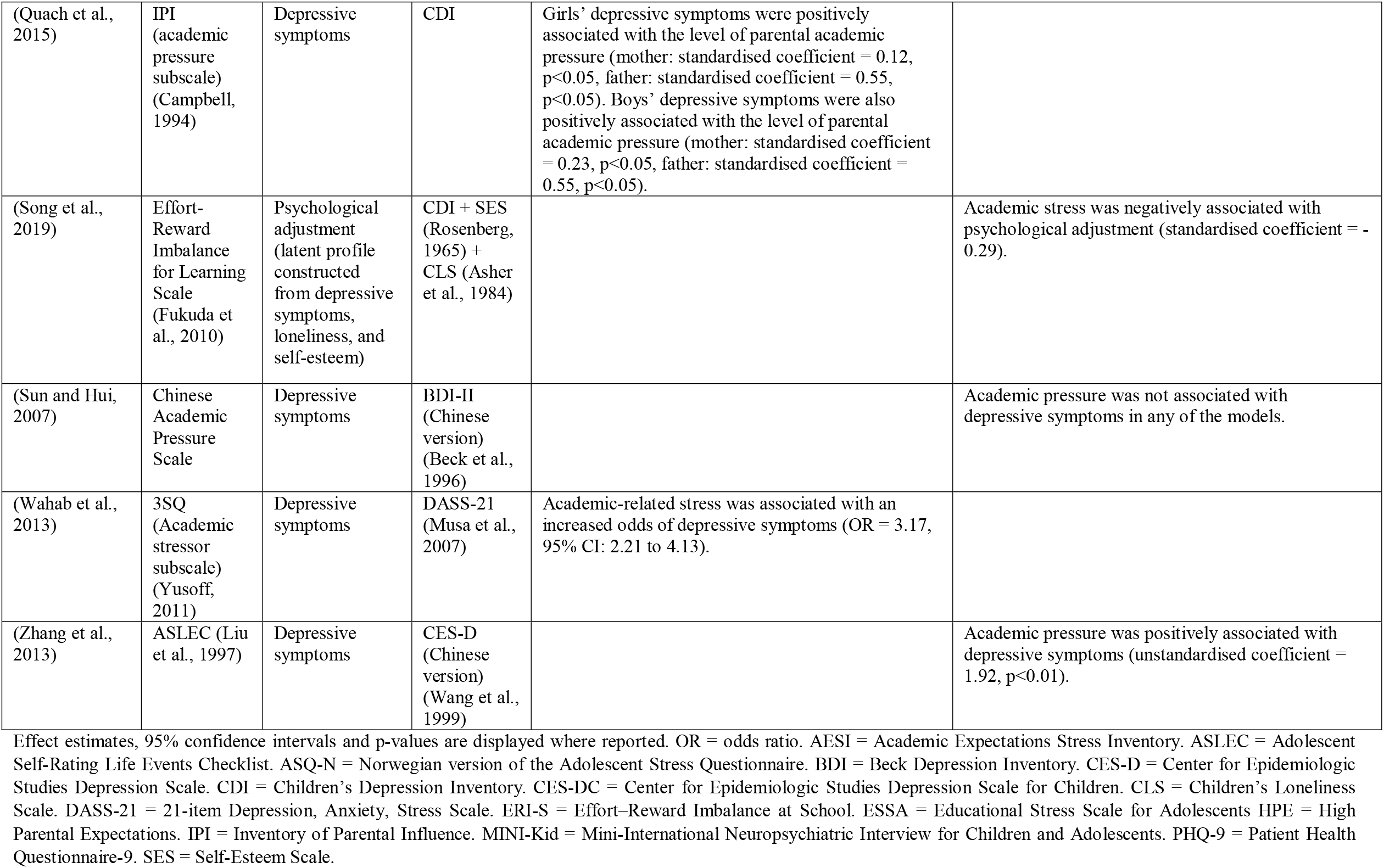
Summary of studies with depressive symptoms as the outcome.

Of the three prospective cohort studies, two found evidence of an association between academic pressure and depressive symptoms at follow-up after adjusting for depressive symptoms at baseline (Fu et al., 2022; Kaman et al., 2021). In a study of 632 German adolescents, academic pressure was positively associated with depressive symptoms at the 5-year follow-up among boys but not girls (Kaman et al., 2021). An analysis of the first two waves of the China Education Panel survey found that academic stress at baseline was positively associated with depressive symptoms at follow-up one year later (Fu et al., 2022). In a study of 520 English adolescents, there was no evidence that depressive symptoms increased as examinations approached (6 to 8 weeks before examinations compared to one week before) (Locker and Cropley, 2004).

#### Anxiety

Five cross-sectional studies reported anxiety outcomes (Table 3). In all cross-sectional studies, academic pressure was found to be positively associated with anxiety symptoms. In the single prospective cohort study, there was no evidence that students’ level of anxiety symptoms were associated with proximity to examinations (Locker and Cropley, 2004).

**Table 3.** Summary of anxiety symptom outcomes.

| Reference | Academic pressure measure | Outcome | Measure | Results summary |  |
| --- | --- | --- | --- | --- | --- |
|  |  |  |  | Unadjusted | Adjusted |
| (Li et al., 2022)/(Jiang et al., 2021) | Unnamed 4-item scale | Anxiety symptoms | Brief Symptom Inventory (Anxiety Subscale) (Derogatis and Spencer, 1993) | Academic pressure was positively associated with anxiety symptoms (unstandardized coefficient = 0.327, $p < 0.001$) | |
| (Locker and Copley, 2004) | Proximity to exams | Anxiety symptoms | RCMAS (Reynolds and Richmond, 1978) | No change in levels of anxiety occurred between the two times of testing (6 to 8 weeks prior to exams vs week before exam) |  |
| (Nguyen et al., 2013) | ESSA(Sun et al., 2011) | Anxiety (clinical cut-off) | Unnamed measure (Nguyen et al., 2007) | Students with high educational stress had an increased odds of anxiety in comparison to those with low educational stress (OR = 3.72, 95% CI: 2.56-5.41). Students with medium educational stress had no increase(OR = 1.32, 95% CI: 0.87-1.99). | Students with high educational stress had an increased odds of anxiety in comparison to those with low educational stress (OR = 3.49, 95% CI: 2.39-5.09). Students with medium educational stress had no increase (OR = 1.31, 95% CI: 0.87-2.0). |
| (Quach et al., 2015) | IPI (academic pressure subscale)(Campbell, 1994) | Anxiety symptoms | RCMAS | Girls' anxiety symptoms were positively associated with the level of parental academic pressure (mother: standardised coefficient = 0.08 ($p < 0.05$), father: standardised coefficient = 0.57($p < 0.05$)). Boys' anxiety symptoms were positively associated with academic pressure from their father (standardised coefficient = 0.74 ($p < 0.05$), but not from their mother (standardised coefficient = 0 ($p > 0.05$)). | |
| (Wahab et al., 2013) | 3SQ (Academic related stressor subscale)(Yusoff, 2011) | Anxiety symptoms | DASS-21 (Musa et al., 2007) | Academic-related stress was associated with an increased odds of anxiety symptoms (OR = 2.63, 95% CI: 1.68 -3.57). |  |
| (Wen et al., 2020) | Single academic pressure item | Anxiety symptoms | Mental Health Test (Zhou, 1991) |  | Students who reported very high levels of academic pressure were more likely to have moderate (OR = 6.52, 95% CI: 2.67-16.0) and severe anxiety (OR = 11.58, 95% CI: 4.16-32.3) than those with very low academic stress. Students who reported high levels of academic pressure were more likely to have moderate (OR = 6.12, 95% CI: 2.77-13.51) and severe anxiety (OR = 5.89, 95% CI: 2.28-15.23) than those with very low academic stress. |

Effect estimates, 95% confidence intervals and p-values are displayed where reported. OR = odds ratio. DASS-21 = 21-item Depression, Anxiety, Stress Scale. ESSA = Educational Stress Scale for Adolescents. IPI = Inventory of Parental Influence. RCMAS = Revised Children's Manifest Anxiety Scale. 3SQ = Secondary School Stressor Questionnaire.

#### Mixed depressive and anxiety symptoms

Mixed anxiety and depressive symptoms was the most commonly assessed outcome, with 20 studies included (Table 4). Eight cross-sectional studies reported a positive association between academic pressure and psychosomatic symptoms. In a repeated cross-sectional survey of secondary school students in Sweden, the magnitude of the association between academic pressure and psychosomatic symptoms had increased in recent years (Högberg et al., 2020) Compared with adolescents in 1993, school-related stress had a stronger positive association with psychosomatic symptoms among adolescents in 2017. In the single longitudinal study (of 767 Norwegian adolescents), school-related stress at baseline was found to be positively associated with psychosomatic symptoms at 6- and 12-month follow-ups (Torsheim et al., 2003).

**Table 4.** Summary of mixed anxiety and depressive symptoms outcomes.

| Reference | Academic pressure measure | Outcome | Measure | Results summary |  |
| --- | --- | --- | --- | --- | --- |
|  |  |  |  | Unadjusted | Adjusted |
| (Bersia et al., 2022) | HBSC single schoolwork pressure item | Psychological health complaints (two or more symptoms experienced more than once a week) | HBSC-SCL (psychological subscale) |  | Pressure from schoolwork was associated with increased odds of experiencing two or more psychological health complaints.<br>Males:<br>Age 11: OR = 2.54 (95% CI: 2.3-2.81)<br>Age 13: OR = 2.15 (95% CI: 1.95 – 2.38)<br>Age 15: OR = 2.2 (95% CI: 2.01 – 2.42)<br>Females:<br>Age 11: OR = 2.67 (95% CI: 2.6 – 2.97)<br>Age 13: OR = 2.49 (95% CI: 2.26 – 2.97)<br>Age 15: OR = 2.29 (95% CI: 2.04 – 2.57) |
| (Chyu and Chen, 2022) | Unnamed 17-item scale | Mental distress | Brief Symptom Inventory (Anxiety, Depression & Somatization Subscales) (Derogatis and Spencer, 1993) | Academic stress was positively associated with mental distress (unstandardised coefficient = 0.53, p<0.001) |  |
| (Cosma et al., 2020) | HBSC single schoolwork pressure item | Psychosomatic symptoms | HBSC-SCL |  | Schoolwork pressure had a positive association with psychosomatic health complaints, with a 1-point increase in schoolwork pressure associated with a 1.98-point increase in the frequency of psychosomatic health complaints (95% CI: 1.96-2). |
| (Deb et al., 2015) | Perception about Stress of Adolescents scale | Psychological distress (caseness) | GHQ-28 | Academic stress was associated with binary GHQ scores (OR = 2.3, 95% CI: 1.2 – 4.6) | Academic stress was significantly associated with binary GHQ scores (OR = 2.2, 95% CI: 1.1 – 4.4) |
| (Eriksson and Sellström, 2010) | 3 item HBSC scale | Psychosomatic symptoms | HBSC-SCL |  | A higher level of school demands, was associated with increased odds of psychosomatic symptoms (OR = 1.30, 95% CI: 1.27-1.34). |
| (Haugan et al., 2021) | 3 item HBSC scale | Emotional problems | SDQ (Goodman, 1997) |  | School stress was positively associated with the level of emotional problems for both girls (standardised coefficient = 0.36) and boys (standardised coefficient = 0.39). |
| (Hodge et al., 1997) | 4 items on examination expectations | Psychological distress | GHQ-30 item (Goldberg, 1972) |  | Stress from examination expectations was not associated with psychological distress. |
| (Högberg et al., 2020) | HBSC single schoolwork pressure item | Psychosomatic symptoms | HBSC-SCL |  | School-related stress was positively associated with psychosomatic symptoms. In 1993, a 1-point increase in school-related stress was associated with a 1.4 increase in psychosomatic symptoms. In 2017, a 1-point increase in school-related stress was associated with a 2.2 increase in psychosomatic symptoms (all unstandardised coefficients). |
| (Högberg, 2021) | HBSC single schoolwork pressure item | Psychological health complaints | HBSC-SCL (psychological subscale) | | School stress had a positive association with mental health symptoms in all models ($p < 0.001$). |
| (Hosseinkhani et al., 2020a) | IAASQ (Future Concerns, Academic Competition, and Stress of Parent Involvement subscales) (Hosseinkhani et al., 2020b) | Mental health symptoms | SDQ – Persian version | Academic stress from (a) academic competition and (b) parental involvement was not associated with mental health symptoms (coefficients not reported). Future concerns was positively associated with mental health symptoms (standardised coefficient = 0.12, $p = 0.001$). | Future concerns were positively associated with mental health symptoms (standardised coefficient = 0.08, $p = 0.001$). |
| (Long et al., 2020) | Single perceived exam pressure item | Psychological distress | GHQ-12 (Goldberg and Williams, 1988) | Perceived exam pressure was positively associated with psychological distress (unstandardised coefficient = 0.49, $p < 0.05$). | Perceived exam pressure was positively associated with psychological distress (unstandardised coefficient = 0.47, $p < 0.05$). |
| (Lönnfjörð and Hagquist, 2021) | Single perceived exam pressure item | Psychosomatic symptoms | Psychosomatic Problems Scale (Hagquist, 2001) | Students reporting a high degree (OR = 23.97, 95% CI: 15.72-36.55) or moderate degree (OR = 4.12, 95% CI: 2.89-5.86) of schoolwork pressure had higher odds of reporting a high degree of psychosomatic symptoms in comparison to those with low levels of schoolwork pressure. | Students reporting a high degree (OR = 14.33, 95% CI: 8.72-23.55) or moderate degree (OR = 3.27, 95% CI: 2.16-4.93) of schoolwork pressure had higher odds of reporting a high degree of psychosomatic symptoms in comparison to those with low levels of schoolwork pressure. |
| (Nygren and Hagquist, 2019) | HBSC single school demands item | Psychosomatic symptoms | Psychosomatic Problems Scale | | Students that reported high demands from teachers had higher odds of experiencing a higher degree of psychosomatic problems ($\geq 90^{\text{th}}$ percentile) than students reporting lower levels of teachers demands (OR = 3.76, 95% CI: 3.18-4.45). |
| (Redmond et al., 2022) | HBSC single schoolwork pressure item | Psychosomatic symptoms (two or more symptoms) | HBSC-SCL |  | High schoolwork pressure was positively associated with a high level of psychosomatic symptoms in all three countries (Australia: OR = 3.03 (95% CI= 2.34 – 3.92). England: OR = 3.6 (95% CI= 2.57 – 5.06). Spain: OR = 2.21 (95% CI= 1.57 – 2.86)). |
| (Ringdal et al., 2020) | ASQ-N (school performance subscale) (Moksnes et al., 2010) | Depressive and anxiety symptoms | 10-item Hopkins Symptom Checklist (Strand et al., 2003) | | School-related stress was positively associated with symptoms of anxiety and depression (unstandardised coefficient = 0.18, $p < 0.001$). |
| (Song et al., 2020) | ASLEC (Liu et al., 1997) | Emotional problems | SDQ – Chinese version |  | Students with high study pressure had increased odds of emotional symptoms in comparison to those with low study pressure (OR = 2.79, 95% CI: 2.32–3.35). |
| (Sonmark et al., 2016) | HBSC single schoolwork pressure item | Psychosomatic symptoms | HBSC-SCL | | High schoolwork pressure was positively associated with the frequency of psychosomatic complaints across all tested school years in both French and Swedish samples.<br>France: age 11 unstandardised coefficient = 4.6 ($p < 0.001$), age 13 unstandardised coefficient = 4.42 ($p < 0.001$), age 15 unstandardised coefficient = 4.72 ($p < 0.001$). Sweden: age 11 unstandardised coefficient = 4.26 ($p < 0.001$), age 13 unstandardised coefficient = 4.92 ($p < 0.001$), age 15 unstandardised coefficient = 4.42 ($p < 0.001$). |
| (Torsheim and Wold, 2001) | 3 item HBSC scale | Psychosomatic symptoms | HBSC-SCL | | Academic stress was positively associated with the frequency of psychosomatic complaints (unstandardised coefficient = 0.23, $p < 0.001$). |
| (Torsheim et al., 2003) | HBSC Academic Stress subscale + High Academic Expectations subscale | Psychosomatic symptoms | HBSC-SCL | | School-related stress at baseline was positively associated with psychosomatic symptoms at 6-month (standardised coefficient = 0.09, $p < 0.01$), and at 12-month (standardised coefficient = 0.11, $p < 0.01$) follow-ups. |
| (Zahir Izuan et al., 2018) | ESSA (Sun et al., 2011) | Mental health (categorised as normal, borderline, abnormal) | SDQ | Students with higher levels of educational stress more likely to have poor mental health, than those with medium or low levels of stress ($\chi^2=57$, $p < 0.001$). | Both students with high education stress (OR = 8.18, 95% CI: 4.25–15.75), and medium educational stress (OR = 2.99, 95% CI: 1.53–5.83) were more likely to have poor mental health than those with low levels of educational stress. |

Effect estimates, 95% confidence intervals and p-values are displayed where reported. OR = odds ratio. ASLEC = Adolescent Self-Rating Life Events Checklist. ASQ-N = Norwegian version of the Adolescent Stress Questionnaire. ESSA = Educational Stress Scale for Adolescents. GHQ = General Health Questionnaire. HBSC-SCL = Health Behaviour in School-aged Children symptom checklist. IAASQ = Iranian Adolescents Academic Stress Questionnaire. SDQ = Strength & Difficulties Questionnaire.

A further 10 studies reported a positive association between academic pressure and mixed anxiety and depressive symptoms, measured as mental or psychological distress (n=3), psychological health complaints (n=2), emotional problems (n=2), mental health symptoms (n=2), and depressive and anxiety symptoms (n=1). One study found no association between stress from examination expectations and psychological distress (Hodge et al., 1997).

#### Service use

In a study of Canadian adolescents, mental health-related hospital admissions were found to be highest from January to April, and in October and November and lowest during periods of school closure in July, August and December (Slaunwhite et al., 2019). In a study of stress-related emergency hospital admissions in England, admissions were highest for adolescents during term-time and lowest during school holidays (Blackburn et al., 2021).

#### Non-suicidal self-harm

We found only one single study that measured NSSH (Table 5). In the time-series study of 1,583 adolescents there was evidence that presentations to hospitals in Oxford (UK) for NSSH varied across the year (Hawton et al., 2003). Presentations were lowest during summer and winter school closures (July to September and December). These findings suggest a possible role for school-based stressors although academic pressure cannot be distinguished from other stressors such as bullying. We found no studies that directly assessed the association between academic pressure and non-suicidal self-injury.

**Table 5.** Summary of service use, NSSH, suicide attempts, suicidal ideation and suicide outcomes.

| Reference | Academic pressure measure | Outcome | Measure | Results summary |  |
| --- | --- | --- | --- | --- | --- |
|  |  |  |  | Unadjusted | Adjusted |
| Service use |  |  |  |  |  |
| (Blackburn et al., 2021) | Timing within year | Service use | Stress-related emergency admission hospital |  | Stress-related admissions were highest during term-time, and lowest during school holidays. |
| (Slaunwhite et al., 2019) | Timing within year | Service use | Mental health-related hospital admission | Psychiatric hospital admissions were highest from January through April, and then in November. Admissions were lower in July, August, and December. |  |
| Non-suicidal self-harm |  |  |  |  |  |
| (Hawton et al., 2003) | Timing within year | Non-suicidal self-harm | Presentation for non-suicidal self-harm to a general hospital between 1990 and 2000 | A decrease in the number of presentations for NSSH from July to September, with smaller reductions also seen in April and December. |  |
| Suicidal ideation |  |  |  |  |  |
| (Ang and Huan, 2006a) | AESI (Ang and Huan, 2006b) | Suicidal ideation | SIQ—Junior High School Version (Reynolds, 1988) | Academic stress had a positive association with suicidal ideation (standardised coefficient = 0.22, $p < 0.001$). | |
| (Masood et al., 2018) | ESSA (Sun et al., 2011) | Suicidal ideation | SIQ | Students who screened positive for suicidal ideation expressed greater levels of academic stress than a matched control group ($t=5.17$, $p=0.001$) | Worry about grades was associated with suicidal ideation (standardised coefficient = 0.12, $p < 0.05$), however there was no association between suicidal ideation and other subscales of the ESSA: pressure from study (0.03), despondency (0.12), self-expectation (0.07), and workload (0.02). |
| (Shang et al., 2014) | ERI-S (Li et al., 2010) | Suicidal ideation (previous 6 months) | Single item adapted from the US Youth Risk Behaviour Survey (Zullig et al., 2006) | Effort-reward imbalance was positively associated with suicidal ideation in the previous 6 months (increase in one SD of effort-reward imbalance: OR = 2.34, 95% CI: 1.8-3.05). Effort was positively associated with suicidal ideation in the previous 6 months (increase in one SD of effort-reward imbalance: OR = 1.55, 95% CI: 1.21-1.98). | Effort-reward imbalance was positively associated with suicidal ideation in the previous 6 months (increase in one SD of effort-reward imbalance: OR = 1.77; 95% CI: 1.34-2.35). Effort was positively associated with suicidal ideation in the previous 6 months (increase in one SD of effort-reward imbalance: OR = 1.41, 95% CI: 1.1-1.81). |
| (Sun and Hui, 2007) | Chinese Academic Pressure Scale | Suicidal ideation | Single item in the Chinese-BDI-II (Chinese Behavioral Science Corporation, 2000), and four items from the SSI (Beck et al., 1979) |  | There was no evidence of an association between academic pressure and suicidal ideation. |
| (Zhang et al., 2019) | Single academic pressure item | Suicidal ideation (previous 12 months) | Single item adapted from the Chinese Youth Risk Behaviour Surveillance (Brenner et al., 2002) | For both boys and girls, high levels of academic pressure were associated with suicidal ideation in the past 12 months ($p < 0.001$). | Students with high academic pressure had increased odds of considered suicide in the past 12 months in comparison to those with low academic pressure (Boys: OR = 1.55, 95% CI: 1.30-1.85. Girls: OR: 2.42, 95% CI: 1.05-3). |
| Suicide attempt |  |  |  |  |  |
| (Carbone et al., 2019) | Timing within year | Suicide attempts and ideation | Emergency department attendance related to suicidal ideation or suicide attempt between 2010 and 2014 | Suicides decreased in June, July, August and December in the 5- to 17-year-old group. This trend was not observed in the older age group. |  |
| (Spiller et al., 2020) | Timing within year | Suicide attempts | Records of suicide attempts by self-poisoning from the National Poison Data System database between 2000 and 2018 | There was a decrease in suicide attempts in the non-school months of June, July and August compared to traditional schooling months of September to May for those between the ages of 10 and 18 ($p < 0.01$). This trend was not found in older non-schooling groups. | |
| Suicide |  |  |  |  |  |
| (Hansen and Lang, 2011) | Timing within year | Suicide | Suicides recorded on the Multiple Cause of-Death Public Use Files between 1980 and 2004 | Suicide rates for 14- to 18-year-olds were lower in June, July, August and December, with rates highest in January and March. This trend was not due to the impact of Seasonal Affective Disorder. |  |
| (Lahti et al., 2007) | Timing within year | Suicide | Shooting suicides recorded on official death certificates from 1988 to 2004 in Oulu, Finland | Suicides peaked in the Autumn months of August, September, and October for under 18 year olds ($\chi^2 = 24.7$, $p < 0.001$). This trend was not found in adults. | |
| (Matsubayashi et al., 2016) | Timing within year | Suicide | Death records compiled by the Japanese Ministry of Health, Labour and Welfare between 1974 and 2014 | Increases in suicides for middle and high school students when each school term starts, with the rate of suicide decreased during school breaks. This pattern was not found for university students or working young adults. |  |
| (McCleary et al., 1991) | Timing with year | Suicide | Records of suicide from the National Center for Health Statistics between 1973 and 1985 | In boys under 16 years of age, suicides peaked in January, February, and November. There was a decrease in suicides in June, July and December. This trend was not found in girls. |  |

Effect estimates, 95% confidence intervals and p-values are displayed where reported. OR = odds ratio. AESI = Academic Expectations Stress Inventory. BDI = Beck Depression Inventory. ERI-S = Effort–Reward Imbalance at School. ESSA = Educational Stress Scale for Adolescents. SIQ = Suicidal Ideation Questionnaire. SSI = Scale for Suicidal Ideation.

#### Suicidal ideation

We found five cross-sectional studies that measured suicidal ideation, all finding an association between academic pressure and suicidal ideation (Table 5). Three studies assessed current suicidal ideation. In a study of 200 adolescents from Pakistan, suicidal ideation was positively associated with adolescents’ worries about grades, but there was no evidence of an association with study-related pressure, despondency, self-expectation or level of workload (Masood et al., 2018). Two studies assessed suicidal ideation over the past 6-or 12-months, both finding that academic pressure was associated with suicidal ideation (Shang et al., 2014; Zhang et al., 2019).

#### Suicide attempts

Suicide attempts in adolescents were rarer during non-school months of June, July, and August in two US studies assessing the rate of hospital admissions for suicide attempt across the year (Carbone et al., 2019; Spiller et al., 2020).

#### Suicide

Four studies assessed suicide according to timing within the year. Two studies in the US found adolescent suicide rates were lowest in periods of school closure in the summer and winter (Hansen and Lang, 2011; McCleary et al., 1991) In Japan, suicides also followed a trend where rates were lowest during school closures, and highest at the start of each academic term (Matsubayashi et al., 2016). In a Finnish study of suicides by shooting, a peak was observed in Autumn and there was no clear reduction in suicides during school holidays or examination periods (Lahti et al., 2007).

## Discussion

We found 52 studies investigating the association between academic pressure, or timing within the school year, and mental health outcomes. These studies were from a range of countries, suggesting international concern about the potential mental health consequences of academic pressure. Generally, we found evidence of a positive association between academic pressure and adolescent mental health problems. Of the 52 studies included, 48 found evidence of a positive association with at least one mental health outcome. Most studies assessed mixed anxiety and depression as the outcome, with 19 of the 20 studies finding evidence of a positive association. Positive associations between measures of academic pressure and the mental health outcome of interest were found in 17 of the 19 studies investigating depressive symptoms, five of the six studies investigating anxiety symptoms, and in four of the five studies assessing suicidal ideation. All but one of the nine studies investigating timing with the school year found that suicides, suicide attempts, mental health-related hospital presentations, and non-suicidal self-injury were lowest during periods of school closure, suggesting a potential association with academic pressure within term time.

Most studies used representative samples, and most were relatively large (35 studies had a sample of over 1,000 participants). However, most studies were cross-sectional. This precludes conclusions about the direction of associations as academic pressure may have followed mental health problems. Adolescents with pre-existing mental health problems may be more prone to negative beliefs about school and failure and report higher levels of academic pressure. Many studies insufficiently accounted for variables that might confound associations between academic pressure and mental health. Measures of academic pressure varied in the items they were assessing, and in some cases, measures were single items or unvalidated. We found no studies with samples exclusively from primary schools. This is probably because academic pressure and the risk of mental health problems is higher in older age groups (Solmi et al., 2021; Wuthrich et al., 2020). There were also few studies from low- and lower-middle-income countries. No studies investigated demographic or contextual factors which might modify associations, such as socioeconomic status, area-level deprivation, or the fee-paying status of the school.

Longitudinal studies are needed, to investigate whether academic pressure precedes mental health problems and is a potentially causal hence targetable risk factor. We found three prospective cohort studies that investigated associations between academic pressure and future mental health, after adjusting for baseline mental health (Fu et al., 2022; Kaman et al., 2021; Torsheim et al., 2003). Each found evidence of an association between academic pressure and mental health at follow-up, suggesting potential causal associations. However, these studies had limitations. Adjustment for potential confounders was limited and associations could be explained by unmeasured variables, such as educational attainment or family history of mental health problems. Two of the samples were relatively small compared with other studies in the review (Kaman et al., 2021; Torsheim et al., 2003). Neither study used measures of academic pressure which had evidence of scale development and testing, so measures may not be valid.

Longitudinal studies of timing within the school year focused on suicide, presentations to hospital emergency departments and admissions to mental health hospitals, and tended to consistently show that mental health problems were lowest during school holidays. This suggests that cases of severe adolescent mental health problems are fewer during periods when academic pressure is likely to be lower. However, we cannot rule out other school-related stressors which might explain this association, such as bullying. These were also likely to have included a small number of adolescents not attending school. We did not find any studies reporting the association between timing within the school year with more common adolescent mental health problems, such as depression and anxiety, or with primary care service use where the vast majority of young people present to.

Measures varied in terms of the aspects of academic pressure assessed, whether they were scales or single items. Only a few studies used a measure which had undergone any previous validation. Despite the significant interest in academic pressure, it often remains poorly measured and there appears to be little consensus regarding which are the key contributing constructs. However, findings were generally consistent across measures, indicating that different aspects of academic pressure are associated with adolescent mental health outcomes. Future studies should use scales that measure a broad range of academic pressure domains, but existing scales may need to be adapted to specific countries, cultures, and schooling systems.

Our systematic review has several limitations. The term academic pressure is often used inconsistently across studies (Kaynak et al., 2021). We searched four major databases, carried out forwards and backwards citation searching, and included a broad range of search terms to improve our search strategy. However, it is possible that we missed studies. Due to the diversity of study design and measurement, we were unable to conduct a meta-analysis.

There are concerns about potentially rising levels of academic pressure and the potential impact this may have on adolescent mental health problems. Academic pressure is potentially common and could be modified using whole-school interventions or through longer-term changes to policy. Our findings suggest that academic pressure is a promising potential candidate among modifiable risk factors for adolescent mental health problems. Large-scale longitudinal studies of this topic therefore seem warranted. These studies should adjust for a broad range of confounders and/or use advanced methods of causal inference. If this association is causal, modifying academic pressure through interventions at an individual, school or policy-level could reduce the rising incidence of adolescent mental health problems.

## Supporting information

. Full search terms are available in the appendix.

## Data Availability

All data within this systematic review was extracted from published studies and does not include any individual participant data.

## Acknowledgments

We thank Wenqianglong Li (Ayden Lee) and Jane Hahn for their translation of academic pressure scales.

## Author Contribution

GL, AS & TS designed the study. TS did the search, data extraction, quality assessment, and wrote the paper. GL & CGM contributed to the search and data extraction. TS led the narrative synthesis with input from GL, CGM & AS. All authors contributed to consecutive drafts and approved the final manuscript.

