## Supplementary material for "The association between academic pressure and adolescent mental health problems: A systematic review": . Full search terms are available in the appendix.

Medline (OVID) (between 1946 and Nov 24, 2022)

| 1 | Schools/ |
| --- | --- |
| 2 | Education/ |
| 3 | (educat* or school*).ti,ab,kf,kw. |
| 4 | 1 or 2 or 3 |
| 5 | (pressure* or stress* or worry or worrie? or anxi* adj2 (exam? or examination? or school? or grade* or deadline? or assignment? or mark? or test? or educat* or academic*)).ti,ab,kf,kw. |
| 6 | (pressure* or demand* adj1 (exam? or examination? or school? or teacher? or parent* or academic*)).ti,ab,kf,kw. |
| 7 | ((fail* or pass) adj2 (school* or exam? or examination? or assignment* or mark? or fear* or grade? or test? or coursework)).ti,ab,kf,kw. |
| 8 | academic* adj1 (hardiness* or buoyan* or resilien*).ti,ab,kf,kw. |
| 9 | (school* adj1 (homework*or demand* or burnout*)).ti,ab,kf,kw. |
| 10 | (school work or school-work or schoolwork or high stakes).ti,ab,kf,kw. |
| 11 | (accountab* adj (regime? or system or systems or school? or educat* or teach* or policy or policies)).ti,ab,kf,kw. |
| 12 | 5 or 6 or 7 or 8 or 9 or 10 or 11 |
| 13 | mental disorders/ or anxiety disorders/ or agoraphobia/ or neurotic disorders/ or obsessive-compulsive disorder/ or panic disorder/ or phobic disorders/ or phobia, social/ or mood disorders/ or depressive disorder/ or depressive disorder, major/ or depressive disorder, treatment-resistant/ or dysthymic disorder/ or depression/ or cyclothymic disorder/ or adjustment disorders/ or Mental Health/ or self-injurious behavior/ or self mutilation/ or suicide/ or suicidal ideation/ or suicide, attempted/ |
| 14 | (mental* or psychiatr*) adj2 (health* or ill* or disorder* or diagnos?s or problem*).ti,ab,kf,kw |
| 15 | (mental health services/ or community mental health services/ or community psychiatry/ or Emergency Services, Psychiatric/ or adolescent health services/ or counseling/ or school mental health services/) |
| 16 | (CAMHS or adolescent mental health service*).ti,ab,kf,kw. |
| 17 | (affective disorder* or agoraphobi* or anxiety or GAD or cyclothymi* or depressed or depression or depressive or MDD or mood disorder* or obsessive compulsive disorder* or OCD or panic or parasuicid* or phobi* or adjustment disorder* or dysthymi* or (self adj1 (injur* or harm or mutilat*)) or suicid*).ti,ab,kf,kw. |
| 18 | 13 or 14 or 15 or 16 or 17 |
| 19 | Child/ or Adolescent/ or Students/ |
| 20 | (pupil? or student? or child* or adolescen* or teenager* or youth?).ti,ab,kf,kw. |
| 21 | 19 or 20 |
| 22 | 4 and 12 and 18 and 21 |

PsycINFO (OVID) (between 1806 and Nov 24, 2022)

| 1 | Schools/ |
| --- | --- |
| 2 | Education/ |
| 3 | (educat* or school*).ti,ab,hw,id. |
| 4 | 1 or 2 or 3 |
| 5 | (pressure* or stress* or worry or worrie? or anxi* adj2 (exam? or examination? or school? or grade* or deadline? or assignment? or mark? or test* or educat* or academic*)).ti,ab,hw,id. |
| 6 | pressure* or demand* adj1 (exam? or examination? or school? or teacher? or parent* or academic*)).ti,ab,hw,id. |
| 7 | ((fail* or pass) adj2 (school* or exam? or examination? or assignment* or mark? or fear* or grade? or test? or coursework)).ti,ab,hw,id. |
| 8 | academic* adj1 (hardiness* or buoyan* or resilien*).ti,ab,hw,id. |
| 9 | (school* adj1 (homework*or demand* or burnout*)).ti,ab,hw,id. |
| 10 | (school work or school-work or schoolwork or high stakes).ti,ab,hw,id. |
| 11 | (accountab* adj1 (regime? or system or systems or school? or educat* or teach* or policy or policies)).ti,ab,hw,id. |
| 12 | 5 or 6 or 7 or 8 or 9 or 10 or 11 |
| 13 | mental disorders/ or anxiety disorders/ or agoraphobia/ or obsessive-compulsive disorder/ or panic disorder/ or mood disorders/ or dysthymic disorder/ or depression/ or cyclothymic disorder/ or adjustment disorders/ or Mental Health/ self-injurious behavior/ or self mutilation/ or suicide/ or suicidal ideation/ or suicide, attempted/ |
| 14 | (mental* or psychiatr*) adj2 (health* or ill* or disorder* or diagnos?s or problem*).ti,ab,hw,id. |
| 15 | mental health services/ or community mental health services/ or community psychiatry/ or counseling/ |
| 16 | (emergency psychiatr* or adolescent health service* or school mental health service* or CAMHS or adolescent mental health service*).ti,ab,hw,id. |
| 17 | (affective disorder* or agoraphobi* or anxiety or GAD or cyclothymi* or depressed or depression or depressive or MDD or mood disorder* or obsessive compulsive disorder* or OCD or panic or parasuicid* or phobi* or adjustment disorder* or dysthymi* or (self adj (injur* or harm or mutilat*)) or suicid*).ti,ab,hw,id. |
| 18 | 13 or 14 or 15 or 16 or 17 |
| 19 | Early Adolescence/ or Primary School Students/ or Students/ or Junior High School Students/ or High School Students/ or Middle School Students/ or Primary School Students/ or Intermediate School Students/ or Education Students/ or Students/ or Elementary School Students/ |
| 20 | (pupil? or student? or child* or adolescen* or teenager* or youth?).ti,ab,hw,id. |
| 21 | 19 or 20 |
| 22 | 4 and 12 and 18 and 21 |

ERIC (ProQuest) (between 1966 and Nov 24, 2022)

| 1 | Schools (DE) |
| --- | --- |
| 2 | Education (DE) |
| 3 | School* or educat* |
| 4 | 1 or 2 or 3 |
| 5 | ((pressure* or stress* or worry or worrie? or anxi*) n2 (exam? or examination? or school? or grade* or deadline? or assignment? or mark? or test* or educat* or academic*)) |
| 6 | ((pressure* or demand*) n1 (exam? or examination? or school? or teacher? or parent* or academic*)) |
| 7 | ((fail* or pass) n2 (school* or exam? or examination? or assignment* or mark? or fear* or grade? or test? or coursework)) |
| 8 | academic* n1 (hardiness* or buoyan* or resilien*) |
| 9 | (school* n1 (homework*or demand* or burnout*)) |
| 10 | (“School work” or school-work or schoolwork or “high stakes”) |
| 11 | (accountab* n1 (regime? or system or systems or school? or educat* or teach* or policy or policies)) |
| 12 | 5 or 6 or 7 or 8 or 9 or 10 or 11 |
| 13 | mental disorders/ or anxiety disorders/or depression/ or Mental Health or Self destructive behavior or Suicide (DE) |
| 14 | (mental* or psychiatr*) n2 (health* or ill* or disorder* or diagnos?s or problem*) |
| 15 | Psychiatric services or Mental Health Programs or Psychiatry (DE) |
| 16 | (counsel?ing or “mental health service*” or “emergency psychiatr*” or “community psychiatr*” or “adolescent health service*” or “school mental health service*” or CAMHS or “adolescent mental health service*” |
| 17 | “affective disorder*” or agoraphobi* or anxiety or GAD or cyclothymi* or depressed or depression or depressive or MDD or “mood disorder*” or “obsessive compulsive disorder*” or OCD or panic or parasuicid* or phobi* or dysthymi* or “adjustment disorder*” or (self n1 (injur* or harm or mutilat*)) or suicid*) |
| 18 | 13 or 14 or 15 or 16 or 17 |
| 19 | pupil? or student? or child* or adolescen* or teenager* or youth? |
| 20 | 4 and 12 and 18 and 19 |

Web of Science Core Collection (between 1900 and Nov 24, 2022)

| 1 | TOPIC: (educat* or school*) |
| --- | --- |
| 2 | TOPIC: ((pressure* or stress* or worry or worrie? or anxi*) "NEAR/2" (exam? or examination? or school? Or grade* or deadline? or assignment? or mark? or test* or educat* or academic*)) OR TOPIC: ((pressure* or demand*) "NEAR/1" (exam? or examination? or school? or teacher? or parent* or academic*)) OR TOPIC: ((fail* or pass) "NEAR/2" (school* or exam* or assignment* or mark* or fear* or grade* or test* or coursework)) OR TOPIC: ((academic*) "NEAR/1" (hardiness* or buoyan* or resilien*)) OR TOPIC: (((school*) "NEAR/2" (homework*or demand* or burnout*))) OR TOPIC: (("School work" or school-work or schoolwork or high stakes)) OR TOPIC: (((accountab* ) "NEAR/1" (regime? or system or systems or school? or educat* or teach* or policy or policies))) |
| 3 | TOPIC: ((mental* or psychiatr*) "NEAR/2 "(health* or ill* or disorder* or diagnos?s or problem*)) OR TOPIC: (“mental health service*” or “emergency psychiatr*” or “community psychiatr*” or “adolescent health service*” or “school mental health service*” or CAMHS or “adolescent mental health service*” or counsel?ing) OR TOPIC: ((“affective disorder*” or agoraphobi* or anxiety or GAD or cyclothymi* or depressed or depression or depressive or MDD or “mood disorder*” or “obsessive compulsive disorder*” or OCD or panic or parasuicid* or phobi* or dysthymi* or “adjustment disorder*” or (self NEAR/1 (injur* or harm or mutilat*)) or suicid*)) |
| 4 | TOPIC: (pupil or student* or child* or adolescen* or teenager* or youth*) |
| 5 | 1 and 2 and 3 and 4 |

**List of studies excluded after full-text assessment**

| **First author, year** | **Reason** | **Author communication** |
| --- | --- | --- |
| (Addy et al., 2021) | No exposures of interest |  |
| (Ali et al., 2019) | No outcomes of interest |  |
| (Anniko et al., 2019) | Excluded study design |  |
| (Arthur, 1990) | Full-text inaccessible |  |
| (Arun et al., 2017) | Excluded study design |  |
| (Askell-Williams and Lawson, 2015) | No exposures of interest |  |
| (Assana et al., 2017) | Excluded study design |  |
| (Bashir et al., 2019) | No exposures of interest |  |
| (Beer, 1991) | No exposures of interest |  |
| (Beidel and Turner, 1988) | No exposures of interest |  |
| (Beidel et al., 1994) | No exposures of interest |  |
| (Belhadj Kouider et al., 2015) | Not in English |  |
| (Bourion-Bédès et al., 2022) | No exposures of interest |  |
| (Buddeberg-Fischer et al., 2000) | Not in English |  |
| (Chen, 2018) | Unpublished thesis |  |
| (Chiou et al., 2006) | Excluded study design |  |
| (Cho, 2020) | No exposures of interest | Contacted (no response) |
| (Chusid, 2020) | Unpublished thesis |  |
| (Deng et al., 2022) | Excluded study population |  |
| (Ding, 2002) | Not in English |  |
| (Einstein et al., 2000) | No exposures of interest |  |
| (Elmelid et al., 2015) | No exposures of interest |  |
| (Ertanir et al., 2021) | Excluded study design |  |
| (Fernández-Sogorb et al., 2021) | No outcomes of interest |  |
| (Feurer and Andrews, 2009) | No exposures of interest |  |
| (Friis et al., 2002) | Excluded study population |  |
| (Giota and Gustafsson, 2017) | Excluded study design |  |
| (Govorova et al., 2020) | No outcomes of interest |  |
| (Guiping and Huichang, 2001) | Not in English |  |
| (Högberg et al., 2019) | Excluded study design |  |
| (Högberg et al., 2021) | Excluded study design |  |
| (Hou and Chen, 2016) | Not in English |  |
| (Huang et al., 2009) | No exposures of interest |  |
| (Ishizu, 2017) | No exposures of interest |  |
| (Jerrim, 2021) | No outcomes of interest |  |
| (Julien, 2020) | Unpublished thesis |  |
| (Kaya, 2004) | No exposures of interest |  |
| (Khan et al., 2022) | No outcomes of interest |  |
| (Khanehheshi and Basavarajappa, 2012) | Excluded study design |  |
| (Kim et al., 2018) | No exposures of interest | Contacted (no response) |
| (Kim, 2021) | Excluded study design |  |
| (Kim et al., 2021) | No exposures of interest |  |
| (Koyama et al., 2014) | No exposures of interest |  |
| (Kwon et al., 2015) | No exposures of interest |  |
| (Lazaratou et al., 2010) | No exposures of interest |  |
| (Lee et al., 2006) | No exposures of interest |  |
| (Lee, 2016) | No exposures of interest | Contacted (no response) |
| (Lee et al., 2021) | Excluded study design |  |
| (Li et al., 2021) | Excluded study design |  |
| (Lin and Yusoff, 2013) | Excluded study population |  |
| (Little and Garber, 2004) | No exposures of interest |  |
| (Luo et al., 2020) | Excluded study design |  |
| (Luthar et al., 2021) | No exposures of interest |  |
| (Mardomingo Sanz and Catalina Zamora, 1992) | Not in English |  |
| (Martinez and Bámaca-Colbert, 2019) | No exposures of interest |  |
| (Masi et al., 2000) | No exposures of interest |  |
| (E. M. McMahon et al., 2010) | No exposures of interest |  |
| (Elaine M. McMahon et al., 2010) | No exposures of interest |  |
| (Ming-Yue et al., 2006) | Not in English |  |
| (Mukhopadhyay and Kumar, 1999) | Full-text inaccessible |  |
| (Murberg and Bru, 2004) | No outcomes of interest |  |
| (Natvig et al., 1999) | No exposures of interest | Contacted (no response) |
| (Nguyen and Nguyen, 2019) | No exposures of interest |  |
| (Nonterah et al., 2015) | Excluded study population |  |
| (Okechukwu et al., 2022) | Excluded study population |  |
| (Park and Chung, 2014) | No exposures of interest | Contacted (no response) |
| (Rao, 2008) | Unpublished thesis |  |
| (Rodrigo et al., 2010) | Excluded study design |  |
| (Rodway et al., 2016) | Excluded study design |  |
| (Rodway et al., 2020) | Excluded study design |  |
| (Sandal et al., 2017) | No exposures of interest |  |
| (She et al., 2021) | No exposures of interest |  |
| (Singtakaew and Chaimongkol, 2021) | No exposures of interest |  |
| (Smyth and Banks, 2012) | Excluded study design |  |
| (Stoeber and Rambow, 2007) | Excluded study design |  |
| (Strydom et al., 2012) | Excluded study design |  |
| (Subotic et al., 2008) | Not in English |  |
| (Sund et al., 2003) | Excluded study design |  |
| (Tian et al., 2019) | Full-text inaccessible |  |
| (Tilton-Weaver et al., 2019) | Excluded study design |  |
| (Travers, 2014) | Unpublished thesis |  |
| (Trevethan et al., 2022) | Excluded study design |  |
| (Undheim and Sund, 2005) | No exposures of interest | Contacted (no response) |
| (Victor and Karunakaran, 2018) | No exposures of interest |  |
| (Walburg et al., 2014) | Not in English |  |
| Wang, 2003(Wang, 2003) | Not in English |  |
| (S. Wang et al., 2022) | No outcomes of interest |  |
| (X. Wang et al., 2022) | No exposures of interest |  |
| (Wen et al., 2022) | No exposures of interest |  |
| (Xiang et al., 2019) | No outcomes of interest |  |
| (Xin et al., 2022) | Excluded study population |  |
| (Yadusky-Holahan and Holahan, 1983) | Excluded study design |  |
| (Yan et al., 2018) | Excluded study design |  |
| (Yildirim et al., 2007) | No exposures of interest |  |
| (Yildirim, 2007) | Full-text inaccessible |  |
| (Ying et al., 2020) | No exposures of interest |  |
| (Zakari et al., 2009) | Not in English |  |
| (Zeng et al., 2020) | No outcomes of interest |  |
| (Zhang et al., 2014) | Not in English |  |
| (Zhang et al., 2022) | Excluded study population |  |

**Additional tables**

Table S1. Quality assessment of included studies using the Mixed Methods Appraisal Tool.

| Citation | Representative sample | Appropriate measures | Complete outcome data | Confounders accounted for | Exposure occurred as intended |
| --- | --- | --- | --- | --- | --- |
| Ang & Huan, 2006 | CT | Y | CT | N | Y |
| Bersia et al., 2022 | Y | N | CT | Y | N |
| Blackburn et al., 2021 | Y | Y | Y | Y | Y |
| Carbone, Holzer & Vaughn, 2019 | Y | Y | Y | Y | Y |
| Chyu & Chen, 2022 | N | N | Y | N | Y |
| Cosma et al., 2020 | Y | N | CT | Y | Y |
| Deb, Strodl & Sun, 2015 | CT | N | CT | N | Y |
| Eriksson & Sellström, 2010 | CT | N | Y | Y | Y |
| Fu, Ren & Liang, 2022 | Y | N | CT | N | Y |
| Guo et al., 2014 | CT | Y | Y | Y | Y |
| Hansen & Lang, 2011 | Y | Y | Y | Y | Y |
| Hanspal et al., 2019 | CT | N | CT | N | Y |
| Haugan, Frostad & Mjaavatn, 2021 | Y | N | Y | N | Y |
| Hawton et al., 2003 | Y | Y | Y | Y | Y |
| Ho, Nguyen & Nguyen, 2022 | CT | Y | CT | N | Y |
| Hodge, McCormick & Elliot, 1997 | CT | Y | CT | Y | Y |
| Högberg, Strandh & Haquist, 2020 | Y | N | CT | Y | Y |
| Högberg, 2021 | Y | N | Y | Y | Y |
| Hosseinkhani et al., 2020 | CT | Y | CT | N | Y |
| Huang & Chen, 2015 | Y | Y | Y | N | Y |
| Izuan et al., 2018 | Y | Y | CT | N | Y |
| Jayanthi, Thirunavukarasu & Raajkumar, 2015 | CT | Y | CT | N | Y |
| Jiang, Ren, Jiang & Wang, 2021/Li, Zhang & Cheng, 2022 | Y | N | CT | N | Y |
| Kaman et al., 2021 | CT | N | Y | N | Y |
| Lahti et al., 2007 | Y | Y | Y | Y | Y |
| Liu & Lu, 2011 | CT | Y | CT | N | Y |
| Locker & Cropley, 2004 | CT | Y | N | N | Y |
| Long, Zucca & Sweeting, 2020 | Y | N | Y | Y | Y |
| Lönnfjord & Hagquist, 2020 | Y | N | CT | N | Y |
| Ma, Siu & Tse, 2018 | Y | Y | CT | Y | Y |
| Masood et al., 2018 | CT | Y | CT | Y | Y |
| Matsubayashi, Uedi & Yoshikawa, 2016 | Y | Y | Y | Y | Y |
| McCleary et al., 1991 | Y | Y | Y | Y | Y |
| Moksnes et al., 2016 | CT | Y | CT | N | Y |
| Nguyen et al., 2013 | Y | Y | CT | Y | Y |
| Nygren & Hagquist, 2017 | CT | N | Y | Y | Y |
| Quach et al., 2015 | CT | Y | CT | N | Y |
| Redmond, Garcia-Moya, Moreno, et al., 2022 | Y | N | CT | Y | Y |
| Ringdal et al., 2020 | Y | Y | CT | Y | Y |
| Shang et al., 2014 | Y | Y | CT | Y | Y |
| Slaunwhite et al., 2019 | Y | Y | Y | Y | Y |
| Song, Fu & Wang, 2019 | CT | Y | Y | N | Y |
| Song et al., 2020 | Y | Y | CT | Y | Y |
| Sonmark et al., 2016 | Y | N | Y | N | Y |
| Spiller et al., 2020 | Y | Y | Y | Y | Y |
| Sun & Hui, 2007 | Y | Y | CT | Y | Y |
| Torsheim, Aaroe & Wold, 2003 | CT | N | N | N | Y |
| Torsheim & Wold, 2001 | Y | N | Y | N | Y |
| Wahab et al., 2013 | Y | Y | CT | N | Y |
| Wen et al., 2020 | Y | N | CT | Y | Y |
| Zhang et al., 2013 | CT | Y | CT | N | Y |
| Zhang et al., 2019 | Y | N | Y | Y | Y |

Y = Yes. N = No. CT = Cannot tell.

Table S2. Summary of academic pressure measures used in the included studies.

| **Academic pressure measure** | **Brief description** | **Items** | **Example items** |
| --- | --- | --- | --- |
| Single items | | | |
| HBSC single schoolwork item | Single item on how stressed or pressured respondents feel by their schoolwork, used across HBSC survey years. Responses via a four-point Likert scale. | 1 | “How pressured do you feel by the schoolwork you have to do?” |
| HBSC single school demands item | Single item regarding high teacher demands. Responses via a five-point Likert scale. | 1 | “What characterised schoolwork in your class: Too-high demands from teachers?” |
| Parental pressure single item | Single item regarding parental pressure for good exam performance. | 1 | Not stated |
| Single academic pressure item | Single item regarding stress and pressure from school. Responses via a five-point Likert scale. | 1 | “In the past 12 months, how often did you feel unhappy because of stress/pressure from school?” |
| Single academic pressure item | Single item on the level of academic pressure experienced by respondents. Responses via a five-point Likert scale. | 1 | Not stated |
| Single academic pressure item | Single item on the level of stress experienced by respondents due to academic pressure from parents. Responses via a five-point Likert scale. | 1 | “How stressed do you feel about your parents’ educational expectations?” |
| Single exam pressure item | Single item regarding perceived exam pressure. Responses via a four-point Likert scale. | 1 | “In my school it’s important to pass exams” |
| Scales | | | |
| 3 item HBSC scale | 3-item HBSC academic stress subscale administered across different survey years. Responses are on a five-point scale from strongly disagree (0) to strongly agree (4), with sum score calculated from 0 to 12. Items vary slightly across survey years, but generally concern stress or pressure from schoolwork, difficulty with schoolwork and having too much schoolwork. Unclear if reliability testing or validity assessment has taken place. | 3 | "I have too much schoolwork”  “I am stressed by schoolwork”  “I find schoolwork tiring.” |
| 4 items on Examination Expectations | Factor identified from 27 items related to student's perceptions of support, self-concept, and exam expectations. Features four items relating to perceptions of exams, parental academic pressure, and perceived importance of high exam results for future plans. | 4 | “How important & worthwhile for your future life plans do you rate achieving a good High School Certificate result?”  “What are your parents’ expectations about your High School Certificate result?” |
| 4 items on stress regarding academic achievement | Items focused on stress from homework, achievement, exams and learning, constructed for the studies by Jiang, Ren, Jiang & Wang, (2021) and Li, Zhang & Cheng (2022). Responses via a four-point Likert scale. | 4 | “I am afraid of obtaining a bad grade”  “I feel stress from too much homework”  “I am stressed out from the burden of exams” |
| 17 items on stress from academic expectations | Unnamed scale constructed for Chyu & Chen (2022) study. Features 17 items across four different subscales: stress from parents, stress from teachers, stress from self, and excessive demands. Responses via a five-point Likert scale. | 17 | “I blame myself if I cannot meet my parents’ academic expectations”  “If I have a poor performance in school, I think my teachers are disappointed in me”  “If I cannot meet my own expectations, I am not good enough” |
| Academic stress questionnaire | Chinese measure constructed for the Liu & Lu study (2012). Three items from the homework subscale focus on study pressure and burden. Responses via a five-point Likert scale. | 3 | “To finish my homework makes me feel pressure.”  “My homework is a burden for me.”  “I have a lot of homework to do.” |
| Adapted schoolwork pressure scale | Four items identified from a previous study that measured school-related stress (Murberg & Bru, 2004), focused on demanding schoolwork, a lack of help with schoolwork and concerns with school performance. Responses via a five-point Likert scale. | 4 | “You think that schoolwork has been too demanding”  “You have been concerned about schoolwork you have not done or that you have not done well.” |
| AESI | Validated scale assessing high academic expectations from parents and teachers, and high self-expectations. Responses via a five-point Likert scale. | 9 | “I feel stressed when I know my parents are disappointed in my exam grades.”  “I feel lousy when I cannot live up to my teacher’s expectations.” “When I fail to live up to my own expectations, I feel I am not good enough. “ “When I do not do as well as I could have in an examination or test, I feel stressed.” |
| Chinese Academic Pressure Scale | A Chinese Academic Pressure Scale purposively constructed for the Sun & Hui (2007) study to measure academic stress. Items focus on the importance of good grades, studying effort and competitiveness between peers. | 5 | “It is important to get good grades in our studies”  “Working hard is necessary in getting good grades”  “I feel a sense of competitiveness in my class” |
| Effort-Reward Imbalance for Learning Scale | Chinese scale that estimates effort applied in learning, the level of rewards and positive appraisal for the effort applied and the level of over-investment. An effort-reward imbalance ratio is calculated through a comparison of scores on the effort and reward subscales., with a ratio higher than 1 implying an imbalance and high academic stress. The scale has been validated in both Japan and China. | 10 | “At school, I am often praised by my teachers and parents.” “Because of my efforts and achievements, I will have a bright future.” “When I go to sleep at night, I still think about studying.” |
| ERI-S | Validated questionnaire measuring effort at school, including schoolwork and parental expectations, "reward" addressing academic performance and study prospects, and academic over-commitment. An effort-reward imbalance ratio is calculated through a comparison of scores on the effort and reward subscales, with a ratio higher than 1 implying an imbalance and high academic stress. Responses via a five-point Likert scale. | 19 | Effort (5 items):  “I have too much schoolwork.”  “My parents expect too much of me at school.”  Reward (11 items):  “When I need help, I can get it from my teacher.”  “Most of my schoolmates are kind and helpful.”  Over-commitment (3 items):  “Studying rarely leaves my mind; it is still on my mind when I go to bed.” |
| ESSA | A 16-item validated scale assessing academic stress across 5 domains: pressure from study, workload, worry about grades, self-expectation, and despondency. Responses via a five-point Likert scale. | 16 | Pressure from study (4 items): “I feel a lot of pressure in my daily studying.”  Workload (3 items): “I feel that there are too many tests/exams in the school.”  Worry about grades (3 items): “I feel that I have disappointed my teacher when my test or exam results are not ideal.”  Self-expectation (3 items): “When I fail to live up to my own expectations, I feel I am not good enough.”  Despondency (3 items): “I am very dissatisfied with my academic grades.” |
| HBSC Academic Stress subscale + High Academic Expectations subscale | Composite school-related stress measure comprising items from the HBSC Academic Stress subscale and the High Academic Expectations subscale (Samdal, Wold & Torsheim, 1998). Items focus on stress from academic expectations from teachers and parents, and from schoolwork demands. Responses via a five-point Likert scale. | 5 | "I have too much schoolwork”  “I find schoolwork tiring"  “Teachers expect too much of me in school” |
| HPE | Four-item scale assessing parental academic pressure and expectations. Responses via a five-point Likert scale. | 4 | "My parents expect me to be one of the best students in my class”  "I feel my parents will be disappointed if I don't get very high grades" |
| Perception about Stress of Adolescents | Measure constructed for the study by Deb, Strodl & Sun (2015). Scale features items on the level and sources of academic pressure. Respondents answer either “yes” or “no” to each item. | Unclear | “Do you feel stressed because of academic pressure?” |
| Subscales | | | |
| 3SQ (Academic- related Stressor subscale) | Validated Malay questionnaire designed to perceive sources of stress among adolescents across 6 different domains. The academic related stressor subscale comprises 10 items focused on stress experienced by exams, high workload, high self-expectations, competitive school environment, and difficulties with schoolwork. | 10 | Does it cause you to feel stressful?:  “Examinations.”  “Too much content to learn.”  “Lack of time to do revision.”  “High self-expectation.”  “Competitive learning environment. “ |
| ASLEC  (Study pressure subscale) | Subscale of a validated Chinese measure of negative life events. Items reflect the frequency of negative academic stressors and events in the past 12 months. Negative academic events include exam failure, high workload, and high academic pressure. Respondents indicate the perceived stressfulness of each event through a 5-point Likert scale. | 4 | Experienced:  “Examination failure” “Academic pressure” |
| ASQ-N (school performance subscale) | Norwegian version of the validated Adolescent Stress Questionnaire featuring 9 separate subscales. Academic pressure is measured through items that comprise the "stress of school performance" subscale. Items focus on high school demands and teacher expectations. Items are rated on a five-point Likert scale. | 5 | “Having to study things you don’t understand” “Teachers expecting too much from you”  “Having to concentrate too long during school hours” |
| IAASQ (Future Concerns, Academic Competition, and Stress of Parent Involvement subscales) | A validated 57-item scale assessing academic stress across 9 domains, with 3 subscales relevant to the concept of academic pressure: future concerns, academic competition, and parental involvement. Responses via a five-point Likert scale. | 57 | Future concerns (6 items):  “Worry about failing the university entrance exam.”  “Worry about the future because unemployed people are in society.”  Academic competition (5 items):  “Worry about the behaviour of your friends and peers, in the case of educational failure.”  “Worry about the behaviour of teachers, deputies, and school principals, in the case of educational failure.”  Parent involvement (4 items):  “Parent´s blame you for your low grades.”  “Parent´s compare your grades with your friends or other family members.” |
| IPI (Academic Pressure subscale) | Subscale of the IPI used to assess respondents’ perceptions of parental academic expectations and pressure to achieve. Responses via a five-point Likert scale. | 13 | “School would be more pleasant if my parents were not as strict.”  “When it comes to school, my parents expect the impossible.”  “My parents are pleased only when I get 100% on tests. “ |

AESI = Academic Expectations Stress Inventory. ASLEC = Adolescent Self-Rating Life Events Checklist. ASQ-N = Norwegian version of the Adolescent Stress Questionnaire. ERI-S = Effort–Reward Imbalance at School. ESSA = Educational Stress Scale for Adolescents. HBSC = Health Behaviour in School-aged Children. HPE = High Parental Expectations. IAASQ = Iranian Adolescents Academic Stress Questionnaire. IPI = Inventory of Parental Influence. 3SQ = Secondary School Stressor Questionnaire.
